# ADHD in adults with recurrent depression

**DOI:** 10.1101/2021.02.05.21251211

**Authors:** Victoria Powell, Sharifah Shameem Agha, Rhys Bevan Jones, Olga Eyre, Anita Thapar, Frances Rice

## Abstract

**Background:** Depression is highly heterogeneous in its clinical presentation. Those with attention deficit/hyperactivity disorder (ADHD) may be at risk of a more chronic and impairing depression compared to those with depression alone according to studies of young people. However, no studies to date have examined ADHD in recurrently depressed adults in mid-life.

**Aims:** To investigate ADHD in women in mid-life with a history of recurrent depression.

**Method:** In a sample of women in mid-life (n=148) taken from a UK based prospective cohort of adults with a history of recurrent depression, we investigated the prevalence of ADHD and the association of ADHD with clinical features of depression.

**Results:** 12.8% of the recurrently depressed women had elevated ADHD symptoms and 3.4% met DSM-5 diagnostic criteria for ADHD. None of the women reported having a diagnosis of ADHD from a medical professional. ADHD was associated with earlier age of depression onset, higher depression associated impairment, a greater recurrence of depressive episodes and increased persistence of subthreshold depression symptoms over the study period, higher levels of irritability and increased risk of self-harm or suicide attempt. ADHD symptoms were associated with increased risk of hospitalisation and receiving non-first line antidepressant medication.

**Conclusions:** Higher ADHD symptoms appear to index a worse clinical presentation for depression. Findings suggest that in women with early onset, impairing and recurrent depression, the possibility of underlying ADHD masked by depression needs to be considered.

## Introduction

Depression is a highly heterogeneous disorder that varies in its origins and clinical presentation (1–3). Individuals with attention deficit/hyperactivity disorder (ADHD), a neurodevelopmental disorder, are at high risk of early-onset, recurrent depression (4,5). Epidemiological and clinical studies report associations between early onset depression and neurodevelopmental disorders and traits, especially ADHD (5–9). One four-year follow-up study of depressed adults and community controls found increased odds of having probable ADHD in those with longer lasting, more severe depressive episodes and in those with a reported age of onset of before 21 years old compared to onset after 21 (9). In addition, some studies have found evidence to suggest that there is a neurodevelopmental genetic contribution to some forms of depression, including ADHD and schizophrenia genetic risk (8,10). Those with depression that is accompanied by ADHD are a clinically important group as they show higher rates of antidepressant treatment resistance, suicide and psychiatric hospitalisation compared to those with depression alone (5,11). However, neurodevelopmental disorders that were not identified in childhood can be missed in adults, especially among women (12). There also is emerging evidence to suggest that recurrent adult depression may mask underlying neurodevelopmental disorders including ADHD that are missed in clinical practice (13). However, the rate of ADHD in middle-aged adults with recurrent depression in the community and the effect of ADHD on depression presentation in this group are unknown. In this UK longitudinal study spanning 13 years, we aim to investigate ADHD in women with recurrent depression. First, we investigate the prevalence of ADHD in this group. Then we investigate the clinical features of depression associated with ADHD including age of onset, severity, episode recurrence, subthreshold symptom persistence, suicide and self-harm attempts, psychotic affective symptoms and irritability symptoms. Additionally, we investigate the association of ADHD symptoms with hospitalisation and use of first-line versus non-first-line antidepressant medication, as an indicator of poor treatment response or a complex clinical presentation.

## Method

### Sample

Data came from the Early Prediction of Adolescent Depression (EPAD) study – a prospective longitudinal study of recurrently depressed parents and their offspring based in the UK (14). The baseline sample included 337 parents with recurrent unipolar depression (two or more lifetime DSM-IV Major Depressive Disorder (MDD) episodes confirmed at interview) and their offspring. Families were assessed at 4 time points between April 2007 and September 2020 via interview and questionnaire. The average length of follow up was 16 months between the first and second waves, 13 months between the second and third, and 8 years between the third and fourth. The current study focuses primarily on the fourth assessment wave, as adult ADHD data was collected during this wave. The fourth wave was carried out between 2018 and 2020 and 197 families participated. Study design and participation rates are summarised in Fig 1. Reasons for the lower retention rate between the third and fourth assessment waves include: death of participants, loss to follow-up and withdrawal from the study. Of the 197 participating families at wave 4, 159 mothers had data on their own ADHD symptoms. Of these 159, 148 women (mean age: 53, range: 42 – 67) with complete data on their own ADHD symptoms, clinical features of their depression, irritability and sociodemographic variables formed the primary sample. The authors assert that all procedures contributing to this work comply with the ethical standards of the relevant national and institutional committees on human experimentation and with the Helsinki Declaration of 1975, as revised in 2008. Ethical approval was granted by the Multi-Centre Research Ethics Committee for Wales and from the School of Medicine Ethics Committee, Cardiff University. Written informed consent and assent was gained from each of the participants at each wave. More detailed information about recruitment, assessments and sample characteristics at assessment waves 1 to 3 can be found elsewhere (14,15).

**Fig 1.**
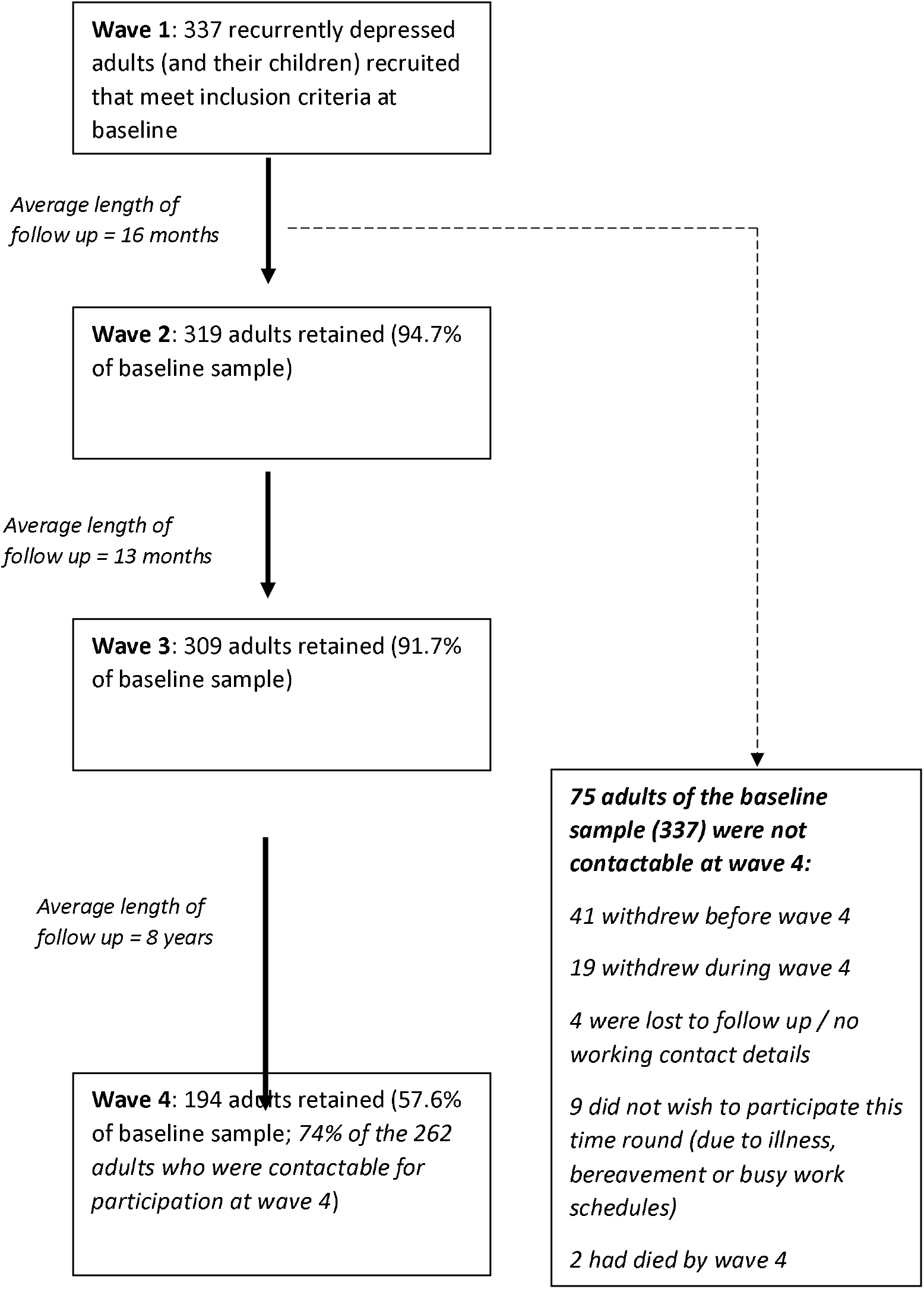
Study design and participation rates across assessment waves of the EPAD study. The Early Prediction of Adolescent Depression (EPAD) study took place over four assessment waves between April 2007 and September 2020 via interview and questionnaire. Numbers of participants reported are those participating at each wave via questionnaire, interview or both. Only 262 adults were contactable at wave 4. Reasons for this include loss of up to date contact details, withdrawal from the study, death and declining to participate due to ill health, bereavement or other commitments such as work (n=75). Of the 262 contactable participants at wave 4, 68 were unresponsive despite multiple communication attempts.

### Overview of assessment procedure

The sample was recruited mainly from general practice surgeries in South Wales (78%). Additional participants were recruited using a database of individuals who had been identified as having previous unipolar depression, via community mental health teams and advertisements in local media and primary care centres (14). The original aim of the study was to conduct a cross-generational study to examine the offspring of parents with recurrent depression, so all participants had biologically related offspring living at home in the age range of 9-17 years at the time of initial recruitment. Recruited adults were screened over the telephone to ensure they met inclusion criteria: suffered from re-current unipolar depression (at least 2 episodes, which were later confirmed using diagnostic interview). Adults were not required to be experiencing a depressive episode at the time of recruitment. Adults who had a lifetime diagnosis of bipolar or psychotic disorder and those who met criteria for DSM-IV mania or hypomania at the time of the interview were excluded. However, adults were still included if they had psychotic or manic experiences without a disorder.

At each assessment wave (1 through 4), the Schedule for Clinical Assessment (SCAN) (16) was used to assess adult DSM-IV MDD based on the symptoms and impairment reported in the preceding month. All cases meeting criteria for diagnosis in addition to subthreshold cases were reviewed by two psychiatrists and diagnoses were made according to clinical consensus. At the baseline assessment, participants were additionally asked to report on their worst ever and second worst ever episodes of depression and associated impairment. They were also asked to retrospectively report on various clinical features of their depression including first age of onset. A life history calendar approach was used for all retrospectively reported information to aid recall (17). At assessment waves 2 through 4, participants were asked to report on episodes of depression they had experienced since the previous assessment wave and to report on associated impairment. Participants additionally completed a questionnaire booklet which included questions relating to sociodemographic information, family structure and relationships, and psychiatric symptom measures such as the Beck Depression Inventory (BDI) (18).

## Measures

### Adult ADHD

Participants completed the Adult ADHD Investigator Symptom Rating Scale (AISRS) (19) via questionnaire at wave 4. The AISRS consists of 18 items based on the DSM-5 (20) symptoms of ADHD. Symptoms were rated on a 4-point scale of “Never/Rarely” (0), “Sometimes” (1), “Often” (2) and “Very often” (3).

Predictor variables were 1) a total ADHD symptom score (possible range: 0-54) and 2) a binary variable of “elevated ADHD symptoms”. Those meeting the AISRS clinical cut point by scoring 24 or higher (21) were classed as having elevated ADHD symptoms.

Diagnosis: Participants were also asked to complete a further set of questions in the questionnaire if they reported ADHD symptoms: at what age they could recall these symptoms starting in approximate years and to indicate whether the symptoms impacted on functioning in a number of areas of life. For descriptive purposes, participants were classed as meeting DSM-5 (20) criteria for adult ADHD diagnosis if they reported at least 5 current symptoms, an age of onset of these problems prior to 12 years old and associated impairment in home, work or social life.

During the interview at assessment wave 4, participants were also asked to report on their recent and current service use and any diagnoses they had received from a medical professional using a questionnaire adapted from the Children’s Services Interview (22), which was used to establish if any participants had received a clinical diagnosis of ADHD.

### Clinical Features of Depression

#### Age of onset

Participants were asked at baseline interview to report the age of onset (in years) of their first MDD episode. Age of onset was dichotomised so that onset at 25 years old or earlier was classed as early-onset depression based on previous studies (23).

#### Severity

Impairment associated with depression was measured using the Global Assessment of Functioning (GAF) (24), which was assessed for the worst ever episode reported by the participant at baseline interview and at each subsequent assessment phase for current/recent depression. A binary measure was derived capturing whether participants had ever had a GAF score <= 50 during the study period with GAF scores below or equal to 50 indicating serious impairment in work or social life (24).

#### Episode recurrence

A count variable of the recurrence of depressive episodes across the study period was derived. This included a count of the number of assessment points at which participants met clinical criteria for MDD plus the number of times participants reported being depressed since the last assessment wave (possible range=0-7).

#### Subthreshold persistence

In addition to recurrence of depression episodes meeting diagnostic criteria, chronicity of subthreshold symptoms over time is common and impairing (25,26).

Thus, as an indicator of persistence of depression symptoms over time, self-report questionnaire measures of depressive symptoms at waves 1 through 4 were used to derive a measure of subthreshold depression persistence. These were the Patient Health Questionnaire (PHQ) (27) at waves 1 and 4 and the Beck Depression Inventory (BDI) (18) at waves 2 and 3 – both valid and reliable measures of depressive symptoms (27–29). Depressive symptom scores were calculated at each wave. Those falling into the “intermediate” symptoms group (between the “none/mild” and “severe” group: scoring 5-19 on the PHQ or 10-29 on the BDI) were defined at each wave based on the cut points validated for the PHQ and BDI (27,28). A count of the number of times symptoms were of the intermediate level was derived (possible range=0-4).

#### Suicide and self-harm attempt

The SCAN (16) was used to assess self-harm or suicide at each wave with the question “Have you thought about harming yourself or even made an attempt at suicide during the last month?”. Participants responses were coded as ‘absent’ (0), ‘intrusive thoughts but no attempt’ (1), ‘injured self but no serious harm resulted’ (2), ‘injured self and serious harm resulted’ (3) or ‘made an attempt at suicide designed to result in death’ (4). A binary variable capturing whether participants had ever reported self-harm or suicide attempts at any of the four assessment waves was derived.

#### Psychotic affective symptoms

A binary variable capturing whether participants had ever endorsed psychotic affective symptoms during the SCAN (16) at waves 1 to 4 was derived. Psychotic affective symptoms assessed were delusions of guilt, delusions of catastrophe, hypochondriacal delusions and auditory hallucinations.

#### Irritability

Participants completed the Affective Reactivity Index (ARI) (30) - a valid, reliable measure of irritability – via questionnaire at wave 4. Though it was developed to assess irritability in children and adolescents, it has been shown to be a reliable and valid measure in adult samples (30). The ARI consists of 7 items with responses of “Not true” (0), “Somewhat true” (1) or “Certainly true” (2), which are summed to give a total score (possible range=0-14). Those scoring higher than 2 were classed as meeting the clinical cut point for irritability for descriptive purposes (31).

#### Hospitalisations

Participants reported the number of times they had ever been hospitalised due to depression at baseline, and the number of times they had been hospitalised since the last assessment wave at waves 2 and 3. At wave 4, participants were not asked explicitly to report the number of hospitalisations, but they were asked to give details of what happened during any depressive episodes experienced since the last wave, which gave an opportunity to disclose hospitalisations. From this information, a binary variable was derived capturing whether participants had ever been hospitalised due to depression.

#### Antidepressant medication

At wave 4, participants reported what medication they were currently taking, including medication for depression. Based upon UK National Institute for Health and Care Excellence (NICE) clinical guidelines for management of adult depression and the British National Formulary (BNF) (32,33), a binary variable of no depression treatment or first-line depression treatment (use of an SSRI) versus non-first-line depression medication (use of an antidepressant other than an SSRI, use of two or more antidepressants, or an antidepressant augmented with lithium or an antipsychotic) was derived. The use of non-first-line antidepressant medication might indicate either poor response to standard, first-line antidepressants (SSRIs), or the need to address complexities in a patient’s depression presentation (32).

### Confounders

All regression analyses presented are adjusted for sociodemographic factors associated with depression that might confound associations (34). The confounders adjusted for are financial status and educational attainment (assessed as having attained qualifications at end of compulsory education, e.g. GCSEs or equivalent qualifications (Yes/No)).

### Analysis

#### Association between ADHD and clinical features of depression

A series of linear and logistic regressions were conducted as appropriate to test the association of ADHD symptoms and clinical features of depression. Continuous ADHD symptoms were standardised so that a one-unit increase was equivalent to a standard deviation increase. All regressions were repeated for ADHD defined as elevated symptoms, entered as a binary variable instead of a continuous variable. This was to test whether the depression presentation of those above the AISRS clinical cut point for ADHD (≥24) was significantly different to those below the cut-point.

#### Missing data

Regression analyses were repeated with Inverse Probability Weights (IPW) applied to assess possible bias arising from potential non-random missing data. IPW is a reliable approach to dealing with missing data, particularly in longitudinal cohorts where participants can have missing data for multiple variables (35). Weights were generated using predictors of missingness from the sample (Supplement 1).

## Results

In the primary sample of recurrently depressed females (n=148), 12.8% (n=19) had elevated ADHD symptoms, indicated by having an AISRS score above the validated clinical cut point (>24), and 3.4% (n=5) met DSM-5 diagnostic criteria for adult ADHD. None of the women in the study however, reported having been diagnosed with ADHD by a clinician. Descriptive statistics are shown in Table 1.

**Table 1.** Descriptive statistics

| Variable | Mean (SD) / % (n) | Range |
| --- | --- | --- |
| ADHD symptoms | 11.19 (10.02) | 0 - 50 |
| Age of onset of depression (% onset ≤25 years) | 26.01 (7.98) / 45.3% (67) | 8 - 46 |
| Ever had GAF score ≤ 50<br>(Severe impairment associated with depression) | 73.0% (108) | Binary: 0=No, 1=Yes |
| Number of MDD episodes during study | 2.42 (1.69) | 0 - 7 |
| Subthreshold depression persistence | 1.97 (1.34) | 0 - 4 |
| Ever hospitalised | 17.6% (26) | Binary: 0=No, 1=Yes |
| Use of non-first-line antidepressants | 20.3% (30) | Binary: 0=No, 1=Yes |
| Ever attempted self-harm or suicide during study | 4.1% (6) | Binary: 0=No, 1=Yes |
| Ever had psychotic affective symptoms during study | 12.8% (19) | Binary: 0=No, 1=Yes |
| Irritability score | 2.65 (2.84) | 0 - 14 |

Descriptive statistics in complete case mothers (n=148). *ADHD* attention-deficit/hyperactivity disorder, *MDD* major depressive disorder, *GAF* Global Assessment of Functioning, *SD* standard deviation

### Association between ADHD and clinical features of depression

ADHD symptoms were associated with an earlier age of depression onset (<= 25 years), and associated severe impairment (GAF<=50) (Table 2). ADHD symptoms were also associated with MDD episode recurrence over the 13-year period of the study, and persistent subthreshold depressive symptoms over the study period. ADHD symptoms were associated with increased risk of reporting self-harm or suicide attempt during the study period, but were not associated with psychotic affective symptoms. ADHD symptoms were associated with higher irritability symptoms.

**Table 2.** Association of adult ADHD symptoms (continuous and dichotomised) and clinical features of depression

| Clinical feature of depression | Association with self-reported adult ADHD symptom score |  | Association with dichotomised self-reported adult ADHD symptoms (above clinical cut-point versus below) |  |
| --- | --- | --- | --- | --- |
|  | b or OR (95% CI) | p-value | b or OR (95% CI) | p-value |
| Depression age of onset of 25 or before | OR=-1.45 (1.02, 2.05) | 0.040 | OR=2.09 (0.74, 5.88) | 0.163 |
| Ever had GAF score ≤ 50<br>(Severe impairment associated with depression) | OR=1.73 (1.08, 2.75) | 0.021 | - - - | - - - |
| Number of MDD episodes during study | b=0.74 (0.51, 0.98) | <0.001 | b=1.93 (1.17, 2.69) | <0.001 |
| Subthreshold depression persistence | b=0.22 (0.01, 0.43) | 0.044 | b=0.29 (-0.368, 0.948) | 0.385 |
| Ever hospitalised | OR=1.94 (1.29, 2.91) | 0.001 | OR=4.58 (1.58, 13.25) | 0.005 |
| Use of non-first-line antidepressants | OR=2.04 (1.35, 3.09) | 0.001 | OR=5.85 (1.99, 17.23) | 0.001 |
| Ever attempted self-harm or suicide during study | OR=3.46 (1.46, 8.21) | 0.005 | - - - | - - - |
| Ever had psychotic affective symptoms during study | OR=0.89 (0.56, 1.40) | 0.603 | OR=0.92 (0.22, 3.98) | 0.916 |
| Irritability score | b=1.54 (1.18, 1.90) | <0.001 | b=3.55 (2.32, 4.78) | <0.001 |

In addition, ADHD symptoms were associated with increased odds of ever being hospitalised and treatment with a non-first-line antidepressant medication which may be indicative of poor response to first-line antidepressants or a complex clinical presentation. The frequencies of different psychotropic medication types used are reported in Supplement 2, which includes depression medication and other neuropsychiatric medications. Of the primary sample (n=148), 63.5% (n=94) reported taking psychotropic medication and 19.6% (n=29) reported taking more than one psychotropic medication.

The same associations were tested with ADHD as a binary variable: those above the AISRS clinical cut-point for ADHD vs. those below the cut-point. Elevated ADHD symptoms were associated with increased recurrence of depressive episodes, increased irritability symptoms, increased risk of hospitalisation and taking non-first-line depression medication. ADHD symptoms above the clinical cut point were also associated with a 2-fold increase in odds of early onset depression, but this did not reach conventional thresholds for significance. The association of elevated ADHD symptoms with severe impairment could not be tested, as all participants with elevated ADHD symptoms had had a GAF score below or equal to 50, and the association with suicide and self-harm attempts could not be tested due to small cell sizes.

Results remained similar with IPW applied (Supplement 3). However, the association between total ADHD symptoms and subthreshold persistence attenuated after IPW was applied.

Descriptive statistics in complete case mothers (n=148). ADHD attention-deficit/hyperactivity disorder, MDD major depressive disorder, GAF Global Assessment of Functioning, SD standard deviation Results in complete case women are shown (n=148). ADHD symptoms were dichotomised using the clinical cut point of the Adult ADHD Investigator Symptom Rating Scale AISRS (>24). All participants above the cut point for ADHD had had a GAF score below 50, so the association of dichotomised ADHD and depression impairment could not be tested due to complete separation issues. The association between dichotomised ADHD and suicide or self-harm attempts could not be tested due to small cell sizes. ADHD attention-deficit/hyperactivity disorder, MDD major depressive disorder, GAF Global Assessment of Functioning, b unstandardized beta, CI confidence interval, OR odds ratio

## Discussion

In a prospective, longitudinal study of recurrently depressed women, we investigated the prevalence of ADHD and the effect of comorbid ADHD on the clinical phenotype of depression. Of this sample, 12.8% were found to have elevated ADHD symptoms and 3.4% met DSM-5 diagnostic criteria for ADHD. These are higher than estimates reported in general population samples of adults (2.5%) (36). None of the participants with ADHD appeared to have been clinically recognised by a medical professional. ADHD is not typically assessed in adult clinical settings. However, those with high ADHD symptoms were more likely to be taking non-first-line antidepressant medication, which might suggest that these individuals have been recognised by clinicians as having a more complex depression presentation or showing resistance to first-line antidepressant treatment (32). ADHD was also associated with earlier age of depression onset, higher depression associated impairment, a greater number of depressive episodes over the study assessment period and increased persistence of subthreshold depression symptoms. ADHD symptoms were also associated with higher levels of irritability, increased odds of self-harm or suicide and increased odds of hospitalisation.

Our findings suggest that a proportion of women in mid-life with a history of early-onset, recurrent depression that has been severely impairing may have undetected ADHD. Previous studies have shown associations of ADHD in childhood and adolescence with more severe depressive outcomes in young adulthood, such as depression recurrence, suicide and hospitalisation (4,5). There is also evidence from a national register-based study that those with ADHD and depression may be at higher risk of antidepressant treatment resistance than those with depression alone (11). A four-year follow-up study of depressed adults and controls found that odds of probable ADHD were higher when depression episodes were longer and when participants reported more severe episodes and an age of onset of depression before 21 (9). However, no studies to date have examined recurrently depressed adults in mid-life.

There is much heterogeneity observed in the presentation of depression (1–3), including in its first age-at-onset and chronicity of illness. Our results suggest that part of this may be accounted for or indexed by comorbid ADHD in some cases. It is possible that some individuals, even if they do not fulfil diagnostic criteria for ADHD, have a more ‘neurodevelopmental type’ of depression characterised by early onset, persistence of symptomatology over time and overlap with neurodevelopmental traits, including ADHD symptoms (6–8). For instance, one study found that an earlier onset, more chronic class of depressive symptoms from childhood to early adulthood (age 18) was associated with more neurodevelopment traits (ADHD, pragmatic language skills and autistic traits) and ADHD genetic risk scores (8). Other studies also suggest that the genetic architecture of earlier onset depression may be more neurodevelopmental in nature and tends to be more strongly associated with neurodevelopmental genetic risk (23,37,38). The present study is important and novel because it examines adults in mid-life with recurrent diagnosed depression from a longitudinal cohort spanning 13 years.

Our findings have important clinical implications. They suggest that in women with early onset, recurrent depression in adulthood, the possibility of depression masking underlying ADHD needs to be considered. Effective treatment of ADHD along with depression could improve functioning and associated depression symptoms (39). Even for those who do not meet diagnostic criteria for ADHD, higher ADHD symptoms appear to index a worse clinical picture. It is possible that they represent a ‘neurodevelopmental type’ of depression presentation, including earlier age of onset, increased severity and recurrence and more prominent irritability. Therefore, they may require more frequent follow-up for management of depression. It will be important for future studies to examine whether this group might respond to different types of depression treatment to those that are typically used for depression such as cognitive behavioural therapy and SSRI antidepressants.

Limitations of this work include the measurement of ADHD that was based on a questionnaire measure completed at one assessment wave. Also in this study we focussed on women with adolescent children who had participated in a longitudinal study of recurrent depression so the findings may not apply to males or other groups, for example hospitalised patients. The rate of ADHD observed may be influenced by specific characteristics of the present sample. Rates of ADHD may be higher in this sample because individuals all had recurrent depression. However, given that ADHD is associated with multiple social and educational impairments, substance misuse and antisocial behaviour, those with comorbid ADHD also may be under-represented in this sample. It is also important to note that although use of non-first-line antidepressant medication can indicate poor response to first-line antidepressants or a complex clinical presentation, there may be other reasons that a non-first-line antidepressant is prescribed, for example, experience of side effects. As is common in longitudinal cohort studies, there was some attrition of the sample over time. Analysis of baseline factors associated with drop-out at wave 4 (Supplement 4), showed that while factors such as lower socioeconomic status and education were associated with attrition, other factors associated with severity of depression and history of depression treatment were not. These included the percentage of the participant’s life that they had been unwell since depression onset and previous non-pharmacological treatment for depression (including talking therapies and electroconvulsive therapy). This might suggest that those with more severe depression or a history of depression treatment may be more likely to stay in the current study, which is likely due to the inclusion criteria of the EPAD study, namely that the participant had experienced at least 2 prior episodes of depression. Nevertheless, to account for any potential bias arising from attrition, analyses were adjusted for variables associated with missingness, helping to address potential bias due to missing data (40). In addition, regression results when inverse probability weights were applied remained similar, suggesting bias due to missingness was minimal (Supplement 3) (35).

To conclude, we found that in a sample of recurrently depressed adults, ADHD was associated with an earlier age of depression onset, higher depression impairment, episode recurrence and subthreshold persistence. ADHD symptoms were also associated with increased risk of suicide or self-harm attempt and increased irritability. ADHD symptoms were associated with increased risk of hospitalisation and receiving non-first-line antidepressant medication, indicating poor response to first-line antidepressant treatment or a complex clinical presentation. Higher ADHD symptoms appear to index a worse clinical presentation for depression. Findings suggest that in women with early onset, recurrent depression, the possibility of underlying ADHD masked by depression needs to be considered.

**Author Affiliations**: Cardiff University, Wales, UK; Cwm Taf Morgannwg University Health Board Health Board, Wales, UK

**Corresponding Author**: Victoria Powell (UK)

## Data Availability

The data that support the findings of this study are available on reasonable request from the corresponding author, Victoria Powell. The data are not publicly available due to their containing information that could compromise the privacy of research participants.

## Declaration of Interests

The authors declare that they have no conflicts of interest.

## Funding

The work was supported by the Medical Research Council (MR/R004609/1). The cohort was established with funding from the Jules Thorn Charitable Trust (JTA/06).

### Acknowledgments

We are very grateful to all the participating families in the Early Prediction of Adolescent Depression study. We thank the GPs and psychiatrists who helped with this study, including Daniel Smith, Judith Allardyce and Robert Potter. We thank all of the assistant psychologists and research assistants who carried out data collection.

## Author Contributions

VP and FR conceptualised the study. VP conducted the analysis and drafted the manuscript. All authors provided critical revision to the manuscript, approved the final version and consented to its publication.

## Notes

### Competing Interest Statement

The authors have declared no competing interest.

### Author Declarations

Ethical approval was granted by the Multi-Centre Research Ethics Committee for Wales and from the School of Medicine Ethics Committee, Cardiff University.

