## Supplementary Files for "ADHD in adults with recurrent depression"

**Supplementary Materials**

**Supplement 1: Inverse Probability Weighting**

To address potential bias caused by non-random missing data, Inverse Probability Weighting (IPW) was conducted (Seaman & White, 2013). This involved weighting the analysis sample by the inverse probability of being missing. Variables measured at the first wave were examined as predictors of missingness from the analysis sample (Supplement 4), including baseline reports of financial income, education, presence of a partner, number of children, anxiety or depression problems and comorbid illness. These formed the missingness model from which the weights were created. Minimal missing data on these predictors were singly imputed as the modal value (all predictors had <13% of values missing). The Hosmer-Lemeshow test showed no indication of poor fit for the missingness model (Hosmer-Lemeshow χ2(8)=12.04, p=0.149). Weights ranged from 1.29 to 8.07. Regression analyses were conducted again with the IPW weights applied to address potential bias caused by missing data. Results remained similar to the unweighted results (Supplement 3).

**Supplement 2:** **Psychotropic medications**

| **Drug category** | **N** | **% of 148** |
| --- | --- | --- |
| Selective serotonin reuptake inhibitors (SSRIs) | 70 | 47.3 |
| Other serotonin reuptake inhibitors (SNRIs or SARIs) | 15 | 10.1 |
| First generation antidepressants (tricyclics or MOAIs) | 11 | 7.4 |
| Atypical antidepressants (tetracyclics) | 4 | 2.7 |
| Antipsychotics | 7 | 4.7 |
| Lithium | 4 | 2.7 |
| Mood stabilisers/anti-epileptic | 5 | 2.7 |
| Anti-anxiety (benzodiazapines, propranolol and buspirone) | 11 | 7.4 |
| Insomnia medication (hypnotics) | 4 | 2.7 |
| Gabapentinoids | 10 | 6.8 |

Psychotropic medication use reported at assessment wave 4 in complete case women (n=148). The dashed horizontal line separates medication types into those used for depression treatment based on NICE guidelines and the BNF (above the line) and other neuropsychiatric medications (below the line).

**Supplement 3:** **Association of adult ADHD symptoms and clinical features of depression with IPW applied**

| **Clinical feature of depression** | **Association with self-reported adult ADHD symptom score** | |
| --- | --- | --- |
|  | **b or OR (95% CI)** | **p-value** |
| Depression age of onset of 25 or before | OR=1.47 (1.05, 2.05) | 0.023 |
| Ever had GAF score ≤ 50  (Severe impairment associated with depression) | OR=1.81 (1.20, 2.75) | 0.005 |
| Number of MDD episodes during study | b=0.72 (0.42, 1.03) | <0.001 |
| Subthreshold depression persistence | b=0.07 (-0.18, 0.32) | 0.594 |
| Ever hospitalised | OR=1.76 (1.14, 2.72) | 0.011 |
| Use of non-first-line antidepressants | OR=2.04 (1.39, 2.98) | <0.001 |
| Ever attempted self-harm or suicide during study | OR=2.88 (1.29, 6.44) | 0.010 |
| Ever had psychotic affective symptoms during study | OR=0.87 (0.48, 1.56) | 0.629 |
| Irritability score | b=1.66 (1.22, 2.10) | <0.001 |

Results in complete case women (n=148) with IPW applied are shown. *IPW* inverse probability weighting, *ADHD*

attention-deficit/hyperactivity disorder, *MDD* major depressive disorder, *GAF* global assessment of functioning,

*b* unstandardized beta, *CI* confidence interval

| **Predictor variable (assessment wave 1)** | **OR** | **95% CI** |  | **p-value** |
| --- | --- | --- | --- | --- |
| Approximate gross family income* | 0.81 | 0.71, 0.92 |  | 0.001 |
| Education (GCSEs or equivalent qualifications)* | 0.50 | 0.27, 0.90 |  | 0.020 |
| Number of children* | 1.37 | 1.07, 1.76 |  | 0.013 |
| Age at birth of index child* | 0.97 | 0.94, 1.01 |  | 0.149 |
| Has a partner living with them* | 0.68 | 0.42, 1.09 |  | 0.109 |
| Current extreme or moderate anxiety or depression*  (questionnaire self-report) | 1.66 | 1.17, 2.36 |  | 0.004 |
| Comorbid illness* | 1.47 | 0.78, 2.76 |  | 0.229 |
| Previous non-pharmacological treatment for depression  (e.g. talking therapies, electroconvulsive shock therapy) | 0.75 | 0.47, 1.19 |  | 0.214 |
| Percentage of life unwell since depression onset | 0.98 | 0.79, 1.20 |  | 0.835 |

**Supplement 4: Prediction of missingness from analysis sample**

Within adults who participated at wave 1 (n=337), logistic regressions between numerous predictor variables and being missing from the analysis sample (n=148) were conducted to establish predictors of attrition in this study. * indicates variables that were included in the final missingness model for inverse probability weighting. *ADHD* attention-deficit hyperactivity disorder, *OR* odds Rati, *CI* confidence interval
